## Supplementary material for "Is there a bilingual advantage in auditory attention among children? A systematic review and meta-analysis of standardized auditory attention tests"

| **Section and Topic** | **Item #** | **Checklist item** | **Location where item is reported** |
| --- | --- | --- | --- |
| **TITLE** | | |  |
| Title | 1 | Identify the report as a systematic review. | Title page |
| **ABSTRACT** | | |  |
| Abstract | 2 | See the PRISMA 2020 for Abstracts checklist. | 2 |
| **INTRODUCTION** | | |  |
| Rationale | 3 | Describe the rationale for the review in the context of existing knowledge. | 3-4 |
| Objectives | 4 | Provide an explicit statement of the objective(s) or question(s) the review addresses. | 5-6 |
| **METHODS** | | |  |
| Eligibility criteria | 5 | Specify the inclusion and exclusion criteria for the review and how studies were grouped for the syntheses. | 7 |
| Information sources | 6 | Specify all databases, registers, websites, organisations, reference lists and other sources searched or consulted to identify studies. Specify the date when each source was last searched or consulted. | 8 |
| Search strategy | 7 | Present the full search strategies for all databases, registers and websites, including any filters and limits used. | 8, S1-3 Tables |
| Selection process | 8 | Specify the methods used to decide whether a study met the inclusion criteria of the review, including how many reviewers screened each record and each report retrieved, whether they worked independently, and if applicable, details of automation tools used in the process. | 8 |
| Data collection process | 9 | Specify the methods used to collect data from reports, including how many reviewers collected data from each report, whether they worked independently, any processes for obtaining or confirming data from study investigators, and if applicable, details of automation tools used in the process. | 8-9 |
| Data items | 10a | List and define all outcomes for which data were sought. Specify whether all results that were compatible with each outcome domain in each study were sought (e.g. for all measures, time points, analyses), and if not, the methods used to decide which results to collect. | 8-9 |
|  | 10b | List and define all other variables for which data were sought (e.g. participant and intervention characteristics, funding sources). Describe any assumptions made about any missing or unclear information. | 8-9 |
| Study risk of bias assessment | 11 | Specify the methods used to assess risk of bias in the included studies, including details of the tool(s) used, how many reviewers assessed each study and whether they worked independently, and if applicable, details of automation tools used in the process. | 8-9 |
| Effect measures | 12 | Specify for each outcome the effect measure(s) (e.g. risk ratio, mean difference) used in the synthesis or presentation of results. | 8-9 |
| Synthesis methods | 13a | Describe the processes used to decide which studies were eligible for each synthesis (e.g. tabulating the study intervention characteristics and comparing against the planned groups for each synthesis (item #5)). | 11 |
|  | 13b | Describe any methods required to prepare the data for presentation or synthesis, such as handling of missing summary statistics, or data conversions. | 10, 17 |
|  | 13c | Describe any methods used to tabulate or visually display results of individual studies and syntheses. | 9 |
|  | 13d | Describe any methods used to synthesize results and provide a rationale for the choice(s). If meta-analysis was performed, describe the model(s), method(s) to identify the presence and extent of statistical heterogeneity, and software package(s) used. | 9-11 |
|  | 13e | Describe any methods used to explore possible causes of heterogeneity among study results (e.g. subgroup analysis, meta-regression). | 10 |
|  | 13f | Describe any sensitivity analyses conducted to assess robustness of the synthesized results. | NA |
| Reporting bias assessment | 14 | Describe any methods used to assess risk of bias due to missing results in a synthesis (arising from reporting biases). | 9 |
| Certainty assessment | 15 | Describe any methods used to assess certainty (or confidence) in the body of evidence for an outcome. | Fig 2 |
| **RESULTS** | | |  |
| Study selection | 16a | Describe the results of the search and selection process, from the number of records identified in the search to the number of studies included in the review, ideally using a flow diagram. | 11, Fig 1 |
|  | 16b | Cite studies that might appear to meet the inclusion criteria, but which were excluded, and explain why they were excluded. | NA |
| Study characteristics | 17 | Cite each included study and present its characteristics. | 12-17, Table 1 |
| Risk of bias in studies | 18 | Present assessments of risk of bias for each included study. | 12-15 |
| Results of individual studies | 19 | For all outcomes, present, for each study: (a) summary statistics for each group (where appropriate) and (b) an effect estimate and its precision (e.g. confidence/credible interval), ideally using structured tables or plots. | Table 1, Fig 2 |
| Results of syntheses | 20a | For each synthesis, briefly summarise the characteristics and risk of bias among contributing studies. | 12-17 |
|  | 20b | Present results of all statistical syntheses conducted. If meta-analysis was done, present for each the summary estimate and its precision (e.g. confidence/credible interval) and measures of statistical heterogeneity. If comparing groups, describe the direction of the effect. | 17-18, S5-12 Tables |
|  | 20c | Present results of all investigations of possible causes of heterogeneity among study results. | 17 |
|  | 20d | Present results of all sensitivity analyses conducted to assess the robustness of the synthesized results. | NA |
| Reporting biases | 21 | Present assessments of risk of bias due to missing results (arising from reporting biases) for each synthesis assessed. | 18 |
| Certainty of evidence | 22 | Present assessments of certainty (or confidence) in the body of evidence for each outcome assessed. | Fig 2 |
| **DISCUSSION** | | |  |
| Discussion | 23a | Provide a general interpretation of the results in the context of other evidence. | 19 |
|  | 23b | Discuss any limitations of the evidence included in the review. | 21 |
|  | 23c | Discuss any limitations of the review processes used. | 18 |
|  | 23d | Discuss implications of the results for practice, policy, and future research. | 21-22 |
| **OTHER INFORMATION** | | |  |
| Registration and protocol | 24a | Provide registration information for the review, including register name and registration number, or state that the review was not registered. | 6 |
|  | 24b | Indicate where the review protocol can be accessed, or state that a protocol was not prepared. | 6 |
|  | 24c | Describe and explain any amendments to information provided at registration or in the protocol. | NA |
| Support | 25 | Describe sources of financial or non-financial support for the review, and the role of the funders or sponsors in the review. | NA |
| Competing interests | 26 | Declare any competing interests of review authors. | In cover letter |
| Availability of data, code and other materials | 27 | Report which of the following are publicly available and where they can be found: template data collection forms; data extracted from included studies; data used for all analyses; analytic code; any other materials used in the review. | Table 1, Fig 1 |

*From:*  Page MJ, McKenzie JE, Bossuyt PM, Boutron I, Hoffmann TC, Mulrow CD, et al. The PRISMA 2020 statement: an updated guideline for reporting systematic reviews. BMJ 2021;372:n71. doi: 10.1136/bmj.n71

For more information, visit: <http://www.prisma-statement.org/>

**S1** **Table. Search terms used in the electronic databases under the concept of “auditory attention”.**

| OVID Medline | OVID PsycInfo | EBSCO CINAHL |
| --- | --- | --- |
| Auditory Cortex/ | exp Auditory Cortex/ or exp Auditory Evoked Potentials/ or exp Auditory Neurons/ or exp Auditory Masking/ | (MH "Auditory Cortex") OR (MH "Evoked Potentials, Auditory+") |
| Auditory Perception/ | exp Auditory Perception/ or exp Auditory Thresholds/ or exp Auditory Stimulation/ | (MH "Auditory Perception+") |
| Acoustic Stimulation/ | exp Acoustics/ | (MH "Acoustic Stimulation") OR (MH "Speech Acoustics") |
| ((auditor* or hear* or listen*) adj3 attention).tw,kf. | ((auditor* or hear* or listen*) adj3 attention).tw. | TI ( ((auditor* or hear* or listen*) N2 attention) ) OR AB ( ((auditor* or hear* orlisten*) N2 attention) ) |
| ((auditor* or hear* or listen*) adj3 perce*).tw,kf. | ((auditor* or hear* or listen*) adj3 perce*).tw. | TI ( ((auditor* or hear* or listen*) N2 perce*) ) OR AB ( ((auditor* or hear* or listen*) N2 perce*) ) |
| ((acoustic* or auditor* or hear* or listen*) adj3 stimul*).tw,kf. | ((acoustic* or auditor* or hear* or listen*) adj3 stimul*).tw. | TI ( ((acoustic* or auditor* or hear* or listen*) N2 stimul*) ) OR AB ( ((acoustic* or auditor* or hear* or listen*) N2 stimul*) ) |
| (alert* adj3 (auditor* or hear* or listen*)).tw,kf. | (alert* adj3 (auditor* or hear* or listen*)).tw. | TI ( (alert* N2 (auditor* or hear* or listen*)) ) OR AB ( (alert* N2 (auditor* or hear* or listen*)) ) |
| (vigilan* adj3 (auditor* or hear* or listen*)).tw,kf. | (vigilan* adj3 (auditor* or hear* or listen*)).tw. | TI ( (vigilan* N2 (auditor* or hear* or listen*)) ) OR AB ( (vigilan* N2 (auditor* or hear* or listen*)) ) |
| (arous* adj3 (auditor* or hear* or listen*)).tw,kf. | (arous* adj3 (auditor* or hear* or listen*)).tw. | TI ( (arous* N2 (auditor* or hear* or listen*)) ) OR AB ( (arous* N2 (auditor* or hear* or listen*)) ) |
| (orient* adj3 (auditor* or hear* or listen*)).tw,kf. | (orient* adj3 (auditor* or hear* or listen*)).tw. | TI ( (orient* N2 (auditor* or hear* or listen*)) ) OR AB ( (orient* N2 (auditor* or hear* or listen*)) ) |
| (selective attention* adj3 (auditor* or hear* or listen*)).tw,kf. | (selective attention* adj3 (auditor* or hear* or listen*)).tw. | TI ( (selective attention* N2 (auditor* or hear* or listen*)) ) OR AB ( (selective attention* N2 (auditor* or hear* or listen*)) ) |
| (sustained attention* adj3 (auditor* or hear* or listen*)).tw,kf. | (sustained attention* adj3 (auditor* or hear* or listen*)).tw. | TI ( (sustained attention* N2 (auditor* or hear* or listen*)) ) OR AB ( (sustained attention* N2 (auditor* or hear* or listen*)) ) |
| (executive control* adj3 (auditor* or hear* or listen*)).tw,kf. | (executive control* adj3 (auditor* or hear* or listen*)).tw. | TI ( (executive control* N2 (auditor* or hear* or listen*)) ) OR AB ( (executive control* N2 (auditor* or hear* or listen*)) ) |
| (executive attention* adj3 (auditor* or hear* or listen*)).tw,kf. | (executive attention* adj3 (auditor* or hear* or listen*)).tw. | TI ( (executive attention* N2 (auditor* or hear* or listen*)) ) OR AB ((executive attention* N2 (auditor* or hear* or listen*)) ) |
| (target detect* adj3 (auditor* or hear* or listen*)).tw,kf. | (target detect* adj3 (auditor* or hear* or listen*)).tw. | TI ( (target detect* N2 (auditor* or hear* or listen*)) ) OR AB ( (target detect* N2 (auditor* or hear* or listen*)) ) |

**S2** **Table. Search terms used in the electronic databases under the concept of “population”.**

| OVID Medline | OVID PsycInfo | EBSCO CINAHL |
| --- | --- | --- |
| exp Child/ | pediatrics/ | (MH "Child Care") OR (MH "Child+") OR (MH "Child Behavior+") OR (MH "Child Day Care") OR (MH "Child Development") |
| exp Infant/ | childhood development/ or child characteristics/ | (MH "Infant Development") |
| Adolescent/ | adolescent development/ | (MH "Adolescence") OR (MH "Adolescent Behavior") OR (MH "Adolescent Development") |
| (child* or infan* or toddler* or newborn* or neonat* or baby or babies or kid* or p?ediat* or boy* or girl* or pre-pubesc* or prepubesc* or adolesc* or pubescen* or juvenile* or teen* or youth*).tw,kf. | (child* or infan* or toddler* or newborn* or neonat* or baby or babies or kid* or pediat* or paediat* or boy* or girl* or pre pubesc* or prepubesc* or adolesc* or pubescen* or juvenile* or teen* or youth*).tw. | TI ( (child* or infan* or toddler* or newborn* or neonat* or baby or babies or kid* or pediat* or paediat* or boy* or girl* or “pre-pubesc*” or prepubesc* or adolesc* or pubescen* or juvenile* or teen* or youth*) ) OR AB ( (child* or infan* or toddler* or newborn* or neonat* or baby or babies or kid* or pediat* or paediat* or boy* or girl* or “pre-pubesc*” or prepubesc* or adolesc* or pubescen* or juvenile* or teen* or youth*) ) |
| (preschool* or pre-school* or kindergar* or school age or nursery school* or (day care* not adult*) or schoolchild* or elementary school* or secondary school* or middle school* or high school*).tw,kf. | (preschool* or pre school* or kindergar* or school age or nursery school* or schoolchild* or elementary school* or secondary school* or middle school* or high school*).tw. | TI ( (preschool* or “pre- school*” or kindergar* or school age or nursery school* or “day care* or schoolchild* or elementary school* or secondary school* or middle school* or high school*) ) OR AB ( (preschool* or “pre- school*” or kindergar* or school age or nursery school* or “day care* or schoolchild* or elementary school* or secondary school* or middle school* or high school*) ) |

**S3** **Table. Search terms used in the electronic databases under the concept of “study types and methods”.**

| OVID Medline | OVID PsycInfo | EBSCO CINAHL |
| --- | --- | --- |
| Cross Sectional Studies/ |  | (MH "Quantitative Studies") OR (MH "Quasi-Experimental Studies") OR (MH "Experimental Studies") OR (MH "Empirical Research") OR (MH "Repeated Measures") |
| Longitudinal Studies/ |  | (MH "Behavioral Research") OR (MH "Case Studies") OR (MH "Comparative Studies") OR (MH "Descriptive Research") OR (MH "Exploratory Research") OR (MH "Evaluation Research") OR (MH "Methodological Research") OR (MH "Multimethod Studies") OR (MH "Physiological Studies") OR (MH "Pilot Studies") OR (MH "Predictive Research") OR (MH "Replication Studies") OR (MH "Secondary Analysis") OR (MH "Validation Studies") |
| Cohort Studies/ |  | (MH "Prospective Studies") OR (MH "Cross Sectional Studies") OR (MH "Observational Methods") |
| cross sectional.ab,ti. | (cohort or longitudinal).ti,ab,id. or cohort study.md. or longitudinal study.md. | TI ( cross sectional or longitudinal or cohort or observ* ) OR AB ( cross sectional or longitudinal or cohort or observ* ) |
| longitudinal.ab,ti. | (cross section* or observ$).ti,ab. |  |
| (cohort adj (study or studies)).ab,ti. |  |  |
| cohort analy$.ab,ti. |  |  |
| (observ$ adj3 (study or studies)).ab,ti. |  |  |
| ((auditor* or hear* or listen* or shadowing) adj3 (task* or test* or paradigm*)).ab,ti. | ((auditor* or hear* or listen* or shadowing) adj3 (task* or test* or paradigm*)).ab,ti. | TI ( ((auditor* or hear* or listen* or shadowing) N2 (task* or test* or paradigm*)) ) OR AB ( ((auditor* or hear* or listen* or shadowing) N2 (task* or test* or paradigm*)) ) |
| (attention* adj3 (task* or test*)).ab,ti. | (attention* adj3 (task* or test*)).ab,ti. | TI ( (attention* N2 (task* or test*))) OR AB((attention* N2 (task* or test*)) ) |
| ((mapping or cueing or monitoring) adj task*).ab,ti. | ((mapping or cueing or monitoring) adj task*).ab,ti. | TI ( ((mapping or cueing or monitoring) task*) ) OR AB ( ((mapping or cueing or monitoring) task*) ) |
| (task-switching paradigm or dual-task or behavioral or behavioural).ab,ti. | (task-switching paradigm or dual-task or behavioral or behavioural).ab,ti. | TI ( (task-switching paradigm or dual-task or behavioral or behavioural) ) OR AB ( (task-switching paradigm or dual-task or behavioral or behavioural)) |
| (electroencephalography or EEG or event-related potential* or ERP* or evoked potential* or magnetic resonance imaging or MRI or fMRI or near-infrared spectroscopy or NIRS or fNIRS or magnetoencephalography or MEG or positron emission tomography or PET or diffusion tensor imaging or DTI or neuroimaging or brain imaging or brain mapping or electrophysiological or oscillation).ab,ti. | (electroencephalography or EEG or event-related potential* or ERP* or evoked potential* or magnetic resonance imaging or MRI or fMRI or near-infrared spectroscopy or NIRS or fNIRS or magnetoencephalography or MEG or positron emission tomography or PET or diffusion tensor imaging or DTI or neuroimaging or brain imaging or brain mapping or electrophysiological or oscillation).ab,ti. | TI ( (electroencephalography  or EEG or event-related potential* or ERP* or evoked potential* or magnetic resonance imaging or MRI or fMRI or near-infrared spectroscopy or NIRS or fNIRS or magnetoencephalography or MEG or positron emission tomography or PET or diffusion tensor imaging or DTI or neuroimaging or brain imaging or brain mapping or electrophysiological or oscillation) ) OR AB ( (electroencephalography or EEG or event-related potential* or ERP* or evoked potential* or magnetic resonance imaging or MRI or fMRI or near-infrared spectroscopy or NIRS or fNIRS or magnetoencephalography or MEG or positron emission tomography or PET or diffusion tensor imaging or DTI or neuroimaging or brain imaging or brain mapping or electrophysiological or oscillation)) |
| (heart rate or HR or eye-tracking or eye tracking or eye movement* or pupil*).ab,ti. | (heart rate or HR or eye-tracking or eye tracking or eye movement* or pupil*).ab,ti. | TI ( (heart rate or HR or eye-tracking or eye tracking or eye movement* or pupil*) ) OR AB ( (heart rate or HR or eye-tracking or eye tracking or eye movement* or pupil*) ) |

**S4 Table. Random-effects meta-analytic model summary.**

| Number of studies combined: k = 20  Number of observations: o = 2115 | | | | |
| --- | --- | --- | --- | --- |
|  | SMD | 95%-CI | t | *p*-value |
| Random effects model | -0.0858 | -0.2702; 0.0985 | -0.97 | 0.3421 |
| Prediction interval |  | -0.7517; 0.5800 |  |  |
| Quantifying heterogeneity: | | | | |
| tau^2^ = 0.0931 [95%-CI: 0.0338; 0.2908]; tau = 0.3051 [95%-CI: 0.1838; 0.5393] | | | | |
| *I*^2^ = 65.8% [95%-CI: 45.1%; 78.6%]; *H* = 1.71 [95%-CI: 1.35; 2.16] | | | | |
| Test of heterogeneity: | | | | |
| *Q* = 55.51, *df* = 19, *p*-value < 0.0001 | | | | |

*k*: number of effect sizes; SMD: standardized mean difference; 95%-CI: 95% confidence interval; tau^2^: estimated amount of residual heterogeneity; tau: square root of estimated tau^2^ value; *I*^2^: residual heterogeneity/unaccounted variability; *H*: the square root of *H*^2^ (i.e., unaccounted variability/sampling variability); *Q*: a weighted sum of squares; *df*: degree of freedom.

**S5 Table. Mixed-effects meta-regression model summary, with test measure as the moderator.**

| Mixed-Effects Model (k = 20; tau^2^ estimator: REML) | | | | | |
| --- | --- | --- | --- | --- | --- |
| tau^2^ = 0.0455 (SE = 0.0311), tau = 0.2133, *I*^2^ = 52.11%, *H*^2^ =2.09, *R*^2^ = 51.13% | | | | | |
| Test of Moderators: *F* (*df*1 = 1, *df*2 = 18) = 9.3759, *p*-value = 0.0067 | | | | | |
| Model Results: | | | | | |
|  | Estimated *g* | Standard Error | *df* | *p*-value | 95%-CI |
| Accuracy | 0.1036 | 0.0943 | 18 | 0.2864 | -0.0945; 0.3017 |
| RTs | -0.4432 | 0.1447 | 18 | 0.0067 ** | -0.7472; -0.1391 |

*R*^2^: amount of heterogeneity accounted for; *g*: effect size.

**p* < 0.05. ***p* < 0.01. ****p* < 0.001.

**S6 Table. Subgroup analysis result, stratified by test measure.**

| Test Measure | *g* | 95%-CI | *I*^2^ | *p* _subgroup_ |
| --- | --- | --- | --- | --- |
| Accuracy | 0.1035 | -0.1158; 0.3229 | 47.6% | 0.0014** |
| RTs | -0.3404 | -0.5696; -0.1112 | 59.1% |  |

**p* < 0.05. ***p* < 0.01. ****p* < 0.001.

**S7 Table. Mixed-effects meta-regression model summary for accuracy studies, with participant age as the moderator.**

| Mixed-Effects Model (k = 12; tau^2^ estimator: ML) | | | | | |
| --- | --- | --- | --- | --- | --- |
| tau^2^ = 0.0128 (SE = 0.0206), tau = 0.1133, *I*^2^ = 21.97%, *H*^2^ =1.28, *R*^2^ = 0.00% | | | | | |
| Test of Moderators: *F* (*df*1 = 1, *df*2 = 10) = 2.7507, *p*-value = 0.1282 | | | | | |
| Model Results: | | | | | |
|  | Estimated *g* | Standard Error | *df* | *p*-value | 95%-CI |
| Intercept | -0.3199 | 0.2691 | 10 | 0.2621 | -0.9195; 0.2798 |
| Participant age | 0.0439 | 0.0265 | 10 | 0.1282 | -0.0151; 0.1030 |

**S8 Table. Mixed-effects meta-regression model summary for accuracy studies, with stimulus type as the moderator.**

| Mixed-Effects Model (k = 12; tau^2^ estimator: ML) | | | | | |
| --- | --- | --- | --- | --- | --- |
| tau^2^ = 0.0000 (SE = 0.0117), tau = 0.0011, *I*^2^ = 0.00%, *H*^2^ =1.00, *R*^2^ = 0.00% | | | | | |
| Test of Moderators: *F* (*df*1 = 1, *df*2 = 10) = 0.0033, *p*-value = 0.9550 | | | | | |
| Model Results: | | | | | |
|  | Estimated *g* | Standard Error | *df* | *p*-value | 95%-CI |
| Linguistic stimuli | 0.1099 | 0.1074 | 10 | 0.3303 | -0.1294; 0.3493 |
| Non-linguistic stimuli | -0.0098 | 0.1691 | 10 | 0.9550 | -0.3865; 0.3670 |

**S9 Table. Mixed-effects meta-regression model summary for accuracy studies, with attention components as the moderator.**

| Mixed-Effects Model (k = 12; tau^2^ estimator: ML) | | | | | |
| --- | --- | --- | --- | --- | --- |
| tau^2^ = 0.0000 (SE = 0.0117), tau = 0.0008, *I*^2^ = 0.00%, *H*^2^ =1.00, *R*^2^ = 0.00% | | | | | |
| Test of Moderators: *F* (*df*1 = 3, *df*2 = 8) = 0.8905, *p*-value = 0.4865 | | | | | |
| Model Results: | | | | | |
|  | Estimated *g* | Standard Error | *df* | *p*-value | 95%-CI |
| Sustained attention | 0.2109 | 0.1140 | 8 | 0.1014 | -0.0519; 0.4737 |
| Executive control | -0.1760 | 0.3059 | 8 | 0.5810 | -0.8814; 0.5295 |
| Selective attention | -0.1360 | 0.1860 | 8 | 0.4855 | -0.5648; 0.2929 |
| Auditory attention overall | -0.4032 | 0.2546 | 8 | 0.1519 | -0.9902; 0.1839 |

**S10 Table. Mixed-effects meta-regression model summary for RT studies, with participant age as the moderator.**

| Mixed-Effects Model (k = 8; tau^2^ estimator: ML) | | | | | |
| --- | --- | --- | --- | --- | --- |
| tau^2^ = 0.0144 (SE = 0.0256), tau = 0.1200, *I*^2^ = 22.57%, *H*^2^ =1.29, *R*^2^ = 63.52% | | | | | |
| Test of Moderators: *F* (*df*1 = 1, *df*2 = 6) = 2.0147, *p*-value = 0.2056 | | | | | |
| Model Results: | | | | | |
|  | Estimated *g* | Standard Error | *df* | *p*-value | 95%-CI |
| Intercept | -0.6882 | 0.2805 | 6 | 0.0496 * | -1.3747; -0.0017 |
| Participant age | 0.0484 | 0.0341 | 6 | 0.2056 | -0.0350; 0.1318 |

**S11 Table. Mixed-effects meta-regression model summary for RT studies, with stimulus type as the moderator.**

| Mixed-Effects Model (k = 8; tau^2^ estimator: ML) | | | | | |
| --- | --- | --- | --- | --- | --- |
| tau^2^ = 0.0392 (SE = 0.0406), tau = 0.1981, *I*^2^ = 50.21%, *H*^2^ =2.01, *R*^2^ = 0.66% | | | | | |
| Test of Moderators: *F* (*df*1 = 1, *df*2 = 6) = 0.4022, *p*-value = 0.5494 | | | | | |
| Model Results: | | | | | |
|  | Estimated *g* | Standard Error | *df* | *p*-value | 95%-CI |
| Linguistic stimuli | -0.2063 | 0.2298 | 6 | 0.4040 | -0.7687; 0.3561 |
| Non-linguistic stimuli | -0.1627 | 0.2566 | 6 | 0.5494 | -0.7907; 0.4652 |

**S12 Table. Mixed-effects meta-regression model summary for RT studies, with attention components as the moderator.**

| Mixed-Effects Model (k = 8; tau^2^ estimator: ML) | | | | | |
| --- | --- | --- | --- | --- | --- |
| tau^2^ = 0.0401 (SE = 0.0411), tau = 0.2003, *I*^2^ = 51.70%, *H*^2^ =2.07, *R*^2^ = 0.00% | | | | | |
| Test of Moderators: *F* (*df*1 = 2, *df*2 = 5) = 0.0631, *p*-value = 0.9396 | | | | | |
| Model Results: | | | | | |
|  | Estimated *g* | Standard Error | *df* | *p*-value | 95%-CI |
| Sustained attention | -0.2589 | 0.2564 | 5 | 0.3590 | -0.9181; 0.4003 |
| Executive control | -0.0756 | 0.3633 | 5 | 0.8433 | -1.0095; 0.8583 |
| Selective attention | -0.1049 | 0.2954 | 5 | 0.7370 | -0.8643; 0.6545 |

**S13 Table. Egger’s regression test result.**

|  | Estimate | Standard Error | *t* value | *p*-value |
| --- | --- | --- | --- | --- |
| Intercept | -0.8770 | 1.0493 | -0.836 | 0.414 |
| X | 0.1116 | 0.2054 | 0.543 | 0.594 |
